# Maternal Biological Aging in Mid to Late Pregnancy and across Four Years Postpartum: Evidence for Postpartum Recovery and Disruption by Subsequent Pregnancy

**DOI:** 10.1101/2025.06.30.25330561

**Authors:** Laura Etzel, Patricia Garrett-Peters, Qiaofeng Ye, Abner T. Apsley, Aaliya Ahamed, John Kozlosky, Chris Chairo, Lorrie Schmid, Victoria K. Lee, Cathi Propper, Roger Mills-Koonce, Sarah J. Short, Idan Shalev

## Abstract

**Background:** Pregnancy involves substantial physiological, metabolic, and immunologic adaptation, which may alter trajectories of maternal biological aging. While emerging evidence suggests that pregnancy may transiently accelerate biological aging followed by partial postpartum recovery, longitudinal studies capturing these dynamics, particularly across successive pregnancies, are limited. This study examined changes in maternal biological aging over nearly four years postpartum and assessed whether subsequent pregnancies disrupted recovery trajectories.

**Methods:** Participants (N = 130; aged 18 to 41 years; 23% non-Hispanic Black, 61% non-Hispanic White) were followed longitudinally across pregnancy and nearly four years postpartum. Biological aging was assessed in saliva at up to three timepoints using four biomarkers: absolute telomere length via qPCR, epigenetic clocks (GrimAge2 and PhenoAge), and pace-of-aging (DunedinPACE). Generalized additive mixed models were used to estimate nonlinear change in biological aging across time, with spline terms differentiating early postpartum recovery and later changes moderated by subsequent pregnancy.

**Results:** In non-interaction models, telomere length was stable in the early (∼ 9 months) postpartum period (b=0.49, SE=0.54, *P*=.36), while there was a trend towards deceleration in GrimAge2 (b = −2.01, SE = 1.06, *P*=.06) and a significant deceleration in pace of aging (DunedinPACE; b = −0.24, SE = .04, *P*<.001). In later postpartum (∼43 months), telomere length declined significantly (b = −0.81, SE = .37, *P*=.029), while both GrimAge2 and pace of aging stabilized. A subsequent pregnancy in the later postpartum period was independently associated with shorter telomere length (b = −0.78, SE = .36, *P*=.032), but not with epigenetic clocks. Time-by-late subsequent pregnancy status interaction models revealed that the acceleration in aging markers during the later postpartum period was more pronounced among women who became pregnant again, particularly for DunedinPACE, where a significant interaction (b = 0.20, SE = .07, *P*=.006) suggested that subsequent pregnancy disrupted the slowed pace of aging observed postpartum. Interaction terms for Time-by-late subsequent pregnancy predicting telomere length and GrimAge2 were directionally consistent with this pattern of slowed recovery but did not reach statistical significance. Associations for PhenoAge were consistent in direction with the other aging indices but did not reach significance in either non-interaction or interaction models.

**Conclusion:** Pregnancy may function as a biological stressor that transiently accelerates maternal aging, while the postpartum period offers a potential window for recovery. However, subsequent pregnancies may disrupt this recovery process, compounding biological aging over time. These findings underscore the importance of postpartum recovery and interpregnancy intervals in shaping maternal aging trajectories and warrant further investigation in larger, more diverse samples with additional metabolic covariates.

## 1. Introduction

Pregnancy is a period of dramatic physiological, metabolic, and immune adaptation, demanding extensive changes across nearly every organ system (Hill & Pickinpaugh, 2008). While these changes are critical for supporting fetal development and parturition, they also place substantial demands on the body of the gestational parent. For some individuals, pregnancy may leave lasting biological imprints that contribute to increased risk for conditions such as cardiovascular disease, metabolic dysfunction, and reduced healthspan (Adank et al., 2020; Hauspurg et al., 2019). Preliminary evidence suggests that biological aging may fluctuate over the course of pregnancy and postpartum (Poganik et al., 2023), but the extent, timing, and permanence of these changes remain unclear. These dynamics raise the possibility that biological aging measures could serve as sensitive indicators of pregnancy’s impact on long-term health.

Biological aging refers to the progressive decline in physiological function and system integrity and is increasingly quantified using molecular biomarkers such as telomere length (TL) and DNA methylation (DNAm)–based clocks, including GrimAge2 (Lu et al., 2019), PhenoAge (Levine et al., 2018), and DunedinPACE (Belsky et al., 2020). Telomeres are protective caps at the ends of linear chromosomes that prevent genetic material from deteriorating; however, they shorten with each cell division, which can serve as a marker of biological aging (Blackburn et al., 2015; López-Otín et al., 2013). Epigenetic clocks estimate biological age by measuring DNAm levels at specific sites throughout the genome, providing insights into the aging process and its acceleration under stress (Horvath & Raj, 2018; Kim et al., 2023). These measures reflect cumulative biological wear and tear that may not align with chronological age; an older biological age than chronological age is classified as ‘accelerated biological aging.’ Emerging evidence from both animal and human studies suggests that the period from pregnancy through postpartum may serve as both a catalyst for accelerated biological aging and, paradoxically, a window for recovery (Poganik et al., 2023; Ross et al., 2020). This dynamic may be driven by intersecting biological processes, including heightened inflammation, oxidative stress, and hormonal shifts during gestation, followed by postpartum immunologic remodeling and metabolic recalibration (Bar et al., 2025). However, cross-sectional evidence supports the hypothesis that repeated pregnancies may impose cumulative physiological costs surpassing the body’s ability to fully recover, reflected in shorter telomeres and advanced epigenetic age persisting past the initial postpartum window. For example, in a cohort of young adult women in the Philippines each additional pregnancy was associated with a modest but additive increase in biological age across multiple measures, with effects persisting after adjusting for socioeconomic and behavioral factors (Ryan et al. 2024). Similarly, drawing on data from over 2,300 U.S. women, another study found that higher parity predicted accelerated PhenoAge; this association was attenuated after adjusting for current BMI, suggesting metabolic status as a key mediator (Kresovich et al. 2019).

Longitudinal work has shown that the accelerated biological aging effects of pregnancy may partially reverse during the postpartum period. Women tracked from mid-pregnancy to 12 months postpartum experienced reductions in both GrimAge2 and PhenoAge estimates, alongside a shift toward younger immune cell profiles, supporting the hypothesis of partial biological recovery after childbirth (Ross et al., 2020). However, recovery appears sensitive to contextual and behavioral factors. For example, women who retained more weight up to a year postpartum had diminished reversal of epigenetic age acceleration (Ross et al., 2020).

Retrospective studies further suggest that biological aging effects linked to pregnancy may persist into midlife, particularly among women with adverse metabolic profiles (Kresovich et al., 2019). Subsequent pregnancies in the postpartum period may also disrupt biological recovery. For instance, in the cohort of young adult women in the Philippines, an increased number of pregnancies during the 4-9 year period between study waves predicted greater increases in epigenetic age, suggesting cumulative biological costs; however these measurements were taken after the subsequent pregnancies, limiting insight into within-pregnancy change (Ryan et al., 2024).

Collectively, extant research supports a dynamic model of the influence of pregnancy on biological aging, wherein pregnancy is linked to a transient acceleration of biological aging, followed by at least partial physiological restoration during the postpartum period. Repeated pregnancies may accumulate incremental aging costs, particularly when recovery is incomplete or interrupted by subsequent pregnancies, but within-individual evidence measuring biological aging during repeated pregnancies in lacking, limiting insights into how successive pregnancies shape aging trajectories over time. The current study aims to address this gap by examining longitudinal trajectories of biological aging in pregnancy and postpartum in a diverse cohort, including sampling within successive pregnancies during follow-up. Using four validated biomarkers of biological age (e.g., telomere length, two epigenetic age clocks, and a pace-of-aging indicator), we tracked biological aging from mid/late-pregnancy through over nearly four years postpartum, one of the longest follow-up periods in this literature to date. We tested whether the transition from pregnancy to postpartum was associated with shifts in biological aging and whether a subsequent pregnancy modified these trajectories. In doing so, we provide novel insight into how pregnancy may function as a biological stress test and a potential inflection point in the aging process.

## 2. Materials and Methods

### 2.1 Study Design and Sample Recruitment

Our analytic sample was drawn from the Brain and Early Experiences (BEE) study, an ongoing prospective longitudinal study investigating the health and development of a diverse cohort of pregnant individuals and their children. All pregnant participants identified as women and will be referred to as such hereafter. Details on the study’s recruitment and data collection procedures are available in Mills-Koonce et al. (2022). In summary, between August 2018 and October 2020, women in their second or third trimester of pregnancy were recruited via advertisements in central North Carolina, USA. Eligibility criteria included (1) carrying a singleton pregnancy, (2) primarily speaking English at home, and (3) residing within a 45-minute radius of the study site. Participants attended a prenatal laboratory session at the Biobehavioral Laboratory within the University of North Carolina (UNC) at Chapel Hill School of Nursing, where biological samples and psychosocial data were collected. The study received approval from the Institutional Review Board of UNC Chapel Hill (#17-1914), and all participants provided informed consent.

Saliva samples were collected at three timepoints: during mid-to late-pregnancy (Time 1; average gestational age in weeks = 29±4 weeks), approximately 9-months postpartum (Time 2; average time elapsed from Time 1 = 9.0±2.4 months), and approximately 43-months postpartum (Time 3; average time elapsed from Time 1 = 43±3.8 months). Of the 233 women who completed the prenatal visit, 113 consented to analysis and provided saliva samples at Time 1, 117 at Time 2, and 88 at Time 3. After conducting DNA quality control for TL and epigenetic assays, 271 TL samples and 210 epigenetic samples across 130 women remained in the final analytic dataset. Eighty participants contributed saliva samples at least twice, while 45 provided samples at all three timepoints (see **Supplemental File S1** for detailed sample breakdown).

### 2.2 Telomere length measurement via qPCR

TL was measured using quantitative polymerase chain reaction (qPCR) on DNA extracted from saliva samples, which were collected through passive drool and processed with the QIAamp DNA Mini Kit (Qiagen). The full guidelines for telomere measurement reporting, as recommended by the Telomere Research Network, are provided in **Supplemental File S2**. In summary, for each sample, two qPCR assays were run using a Rotor-Gene Q thermocycler with an uninterruptible power supply – one to quantify telomere content (T) and another to measure genome copy number (S) via the single-copy gene *IFNB1* (Hastings et al., 2020; O’Callaghan & Fenech, 2011; Wolf et al., 2024). The telomere standards contained known concentrations of 84 base pair double-stranded oligomers with 16 repeats of the telomeric sequence (5’-TTAGGG-3’), while the genome copy number standards consisted of 83 base pair double-stranded oligomers corresponding to the *IFNB1* gene region, flanked by *IFNB1* primers. The ratio of telomere content (T) to genome copy number (S) was used to calculate absolute TL in kilobase pairs.

### 2.3 DNA methylation measurement and epigenetic age clock calculation

To assess DNA methylation across the genome, we used the Infinium Methylation EPIC v2.0 BeadChip (Illumina, San Diego, CA, USA). 500ng of genomic DNA from saliva was treated with sodium bisulfite using the Zymo EZ-96 DNA Methylation Kit™ (Zymo Research, Orange, CA, USA), and 200ng of the bisulfite-treated DNA was then amplified, fragmented, and hybridized onto the EPICv2 array. Methylation analysis occurred in two batches, with two samples from each participant per timepoint placed on the same plate to reduce technical variability. A subset of samples (2.5%) from the first batch was rerun in the second batch to quantify batch effects. IDAT files were read into R statistical software using the read.metharray.exp function in the minfi package. CpG probes with a bead count of less than three in 5% or more samples or with an average detection p-value of >0.05 across all samples were excluded. Prior to calculation of epigenetic clocks, all samples were normalized using the beta-mixture quantile normalization (BMIQ) method. Batch effects were corrected for using control probes from the EPICv2 array. The resulting methylation data were used to calculate two second-generation measures of epigenetic age (GrimAge2 and PhenoAge) and a measure of the pace of aging (DunedinPACE). All epigenetic clocks required CpG probes not included in the EPICv2 array, therefore missing probe values were imputed using average values from publicly available adult saliva DNAm data (GSE130153 and GSE111165) as previously done (Apsley et al., 2025). Clocks were computed using modified code from the methylCIPHER and DunedinPACE packages in R. Epigenetic age acceleration measurements were calculated as the difference between chronological age and epigenetic age estimates. Given prior findings that first generation clocks may not be sensitive measurements during transient stressors – such as illness or pregnancy – that may influence the epigenetic landscape (Poganik 2023), we opted to not include first-generation clocks in our analyses.

To adjust for cell type heterogeneity, we applied the Houseman-ReferenceFree method, which estimates cellular proportions from DNA methylation data without requiring predefined reference profiles (Houseman et al., 2016). To determine the optimal number of cell types for saliva tissue, we extracted the 10,000 most variable CpG probes across samples and computed deviance statistics for different cell type estimates using 500 bootstrapped samples. The number of cell types minimizing deviance was selected at 7 (see **Supplemental File S3** for details).

### 2.4 Assessment of subsequent pregnancy

Maternal pregnancy status was self-reported at each timepoint via a health and medication survey item: “Are you currently pregnant (Yes/No)?”. A pregnancy reported at the Time 3 visit or within the preceding 6 months was coded as a late subsequent pregnancy (N=19; 15% of sample). To distinguish these from pregnancies that occurred earlier in the postpartum period, any pregnancies reported between the Time 2 and 6-months prior to Time 3 windows were categorized as early subsequent pregnancy (N=13; minimum length of time between target pregnancy and subsequent early pregnancy = 7.1 months; maximum length of time = 22 months). Due to lack of biological sampling coverage during this earlier timeframe, early subsequent pregnancy was treated as a covariate rather than examined for interactions with time. No pregnancies were reported prior to or at the Time 2 visit.

### 2.5 Covariates

The following covariates were included in all statistical models to account for sociodemographic and health-related confounders: pre-pregnancy body mass index (BMI), gestational age in days at biological sampling during the initial (Time 0) pregnancy, maternal illness burden, self-reported race/ethnicity, and income-to-needs ratio. Pre-pregnancy BMI was defined as the mother’s self-reported weight before pregnancy divided by height squared (kg/m²). Race/ethnicity was categorized as ‘Black’, ‘White’, or ‘Other’ (including Hispanic and multiracial identities). Income-to-needs ratio was calculated as the ratio of total household income to the federal poverty threshold for household size.

Maternal illness burden was assessed at each timepoint using a standardized Health and Medication Survey. Mothers responded “Yes” or “No” to 14 items assessing current or historical diagnoses of chronic and acute conditions, including: cardiac problems or heart disease, diabetes and gestational diabetes, cancer, asthma, epilepsy or seizures, sickle cell disease, stroke, depression or bipolar disorder, anxiety, attention-deficit hyperactivity disorder (ADHD), schizophrenia or borderline personality disorder, other illness, and other genetic disorder.

Affirmative responses were summed at each timepoint to create a total illness score (range: 0– 14), which was then averaged across timepoints to yield a single indicator of maternal illness burden used in the analyses.

### 2.6 Statistical analysis

All statistical analyses were conducted using R version 4.1.2. To address missing data and improve statistical power, we implemented multiple imputation of covariates only via the mice package using predictive mean matching. To quantify the scope of imputation, we identified the number of unique participants with imputed values for each covariate used in our primary models. Of the 130 participants, pre-pregnancy BMI was imputed for 6 (5%) participants, income-to-needs ratio for 8 (6%) participants, and the average maternal illness burden for 3 (2%) participants.

Demographic differences between women with and without a later subsequent pregnancy were assessed via two-tailed Mann-Whitney U test for continuous variables and two-way Chi-Square tests for dichotomous variables. Bivariate correlations were assessed using the ‘cor’ function in base R to calculate Pearson or Phi coefficients as appropriate. TL was normally distributed with two outliers at the right end of the distribution (see **Supplemental File S4** for Q-Q plot and histogram of original TL values); these values were winsorized to the 95^th^ percentile.

All outcome measures were adjusted for estimated cell proportions, and TL, GrimAge2, and PhenoAge were further adjusted for maternal age at collection using linear models. The residuals from these models, representing age- and cell-adjusted biological aging, served as the primary outcomes in subsequent analyses for TL, GrimAge2, and PhenoAge. For DunedinPACE models, maternal age at collection was included as continuous covariate and grand mean centered to facilitate interpretation and reduce potential nonessential collinearity with time trends. All DunedinPACE models use the cell-adjusted DunedinPACE measure as the primary outcome (see **Supplemental File S5** for correlations among raw and age- and cell-residualized aging outcomes).

We fit linear mixed-effects models using the ‘lme4’ and ‘lmerTest’ packages. Each outcome was modeled separately. Time was modeled using a natural spline with two degrees of freedom using the ns() function from the splines package in R. This approach estimates a nonlinear trajectory of change over time by fitting two spline basis functions, each with its own coefficient. These basis functions allow the rate of change in the outcome to vary flexibly across the time span, without assuming a single linear slope. Models included a random intercept for participant ID to account for within-subject correlation across repeated measures and adjustment for relevant covariates as follows:

> Y_ij_=β_0_+β_1_⋅Spline_1_(Time_ij_)+β_2_⋅Spline_2_(Time_ij_)+γ^⊤^X_ij_+b_0j_+ɛ_ij_

Where:

> Y_ij_: the outcome for observation i in subject j
>
> Time_ij_: time in months at observation i for subject j
>
> Spline_1_(⋅),Spline_2_(⋅): spline basis functions from ns(Time, df = 2)
>
> X_ij_: vector of covariates for observation i, subject j
>
> γ^⊤^X_ij_: combined linear effects of all covariates
>
> b_0j_∼N(0,σ_b_ ^2^): random intercept for subject
>
> j ɛ_ij_∼N(0,σ^2^): residual error term

To test whether biological aging trajectories differed by late subsequent pregnancy status, we included a time-by-‘late subsequent pregnancy’ interaction term in our models with ‘earlier subsequent pregnancy’ included as a covariate. Significance was assessed at *p* < .05. To visualize modeled trajectories, we used the ‘ggeffects’ (Lüdecke, 2018) and ‘emmeans’ (Lenth et al., 2025) packages to extract predicted values and generate line plots across time by late subsequent pregnancy status.

## 3. Results

### 3.1 Sample Descriptives

Of 130 women in the analytic sample, 19 (14.6%) reported a subsequent pregnancy that occurred between 30 and 36 months postpartum (hereafter referred to as the “Late Subsequent Pregnancy” group), while 111 (85.4%) reported no such pregnancy. As shown in **Table 1**, women in the Late Subsequent Pregnancy group were significantly younger at the prenatal visit (*M* = 29.47, *SD* = 3.44) compared to those without a later pregnancy (*M* = 31.56, *SD* = 5.46; *P*=.035). There were no statistically significant differences in pre-pregnancy BMI, income-to-needs ratio, or illness burden between groups.

**Table 1.** Descriptive statistics for analytic sample.

| Variable, mean (SD) | No Later Subsequent Pregnancy (N=111) | Subsequent Later Pregnancy (N=19) | p-val |
| --- | --- | --- | --- |
| Age (years) at Time=0 Prenatal Visit | 31.6 (5.5) | 29.5 (3.4) | 0.034 |
| Pre-pregnancy BMI | 27.9 (6.7) | 25.5 (5.0) | 0.080 |
| Income to Needs Ratio | 3.78 (2.6) | 4.09 (2.1) | 0.56 |
| Averaged Illness Burden | 1.64 (1.4) | 1.32 (1.0) | 0.20 |
| Gestational Age (days) at Time=0 Prenatal Visit | 204 (27) | 203 (29) | 0.89 |
| Primary Race/Ethnicity |  |  |  |
| Black | 30 (27%) | 1 (5%) | 0.06 |
| White | 63 (57%) | 16 (84%) |  |
| Other | 18 (16%) | 2 (11%) |  |
| Early Subsequent Pregnancy | 13 (10%) | 0 (0%) | 0.21 |

There was a non-significant difference in the distribution of race/ethnicity by pregnancy group (*P*=.06), with women identifying as Black being less represented among those with a later subsequent pregnancy (1 of 31; 3.2%) compared to those with no subsequent pregnancy (30 of 111; 27%).

Older maternal age was significantly associated with higher income-to-needs ratios (r = 0.31, *P*<.0001; **Table 2**). Pre-pregnancy BMI was positively associated with illness burden (r = 0.20, *P<.*0001) and was significantly lower among women who experienced early (r = –0.20, *P*<.0001) or late (r = –0.09, *P=*.0005) subsequent pregnancies, indicating that lower BMI was associated with a higher likelihood of subsequent pregnancies. Income-to-needs was inversely associated with illness burden (r = –0.20, *P*< .0001) suggesting that women with fewer economic resources tended to have greater health burdens. Additionally, Black race/ethnicity was positively associated with BMI (r = 0.25, *P*<.0001) and negatively correlated with income-to-needs (r = –0.30, *P*<.0001).

**Table 2.** Correlations among variables. Significant correlations are indicated with symbols: *P<.05, **P<.01, ***P<.001, †P<.10. Outcomes are included in their non-age-adjusted form to demonstrate correlations with age.

|  | TL | GrimAge<br>2 | PhenoAge | DunedinPACE | Age at<br>Prenatal<br>Visit | Gestational<br>Age at<br>Prenatal<br>Visit | Pre-pregnancy<br>BMI | Income-to-needs | Illness<br>Burden | Black<br>Race/<br>Ethnicity | White<br>Race/<br>Ethnicity | Other<br>Race/<br>Ethnicity | Early<br>Subsequent<br>Pregnancy |
| --- | --- | --- | --- | --- | --- | --- | --- | --- | --- | --- | --- | --- | --- |
| TL |  |  |  |  |  |  |  |  |  |  |  |  |  |
| GrimAge2 |  |  |  |  |  |  |  |  |  |  |  |  |  |
| PhenoAge | 0.04 |  |  |  |  |  |  |  |  |  |  |  |  |
| DunedinPACE | 0.13† | 0.17* |  |  |  |  |  |  |  |  |  |  |  |
| Age at Prenatal Visit | -0.05 | 0.55*** | 0.13† | -0.12† |  |  |  |  |  |  |  |  |  |
| Gestational Age at Prenatal Visit | -- | -- | -- | 0.06 | -0.01 |  |  |  |  |  |  |  |  |
| Pre-pregnancy BMI | 0.01 | 0.07 | -0.13† | 0.35*** | 0.03 | -0.08** |  |  |  |  |  |  |  |
| Income-to-needs | 0.01 | 0.29*** | 0.08 | -0.21** | 0.31*** | -0.06* | -0.16*** |  |  |  |  |  |  |
| Illness Burden | -0.12 | -0.27*** | -0.10 | 0.28*** | -0.01 | 0.04 | 0.20*** | -0.20*** |  |  |  |  |  |
| Black Race/Ethnicity | 0.09 | 0.27*** | 0.09 | 0.12† | -0.14** | -0.01 | 0.25*** | -0.30*** | -0.06 |  |  |  |  |
| White Race/Ethnicity | 0.24*** | 0.26*** | 0.08 | -0.13† | 0.19*** | 0.001 | -0.25*** | 0.28*** | 0.11* | -0.61*** |  |  |  |
| Other Race/Ethnicity | -0.15* | -0.18** | -0.11† | 0.03 | -0.09* | 0.007 | 0.01 | 0.001 | -0.07 | -0.38*** | -0.51*** |  |  |
| Early Subsequent Pregnancy | -0.09 | -0.06 | 0.06 | -0.17* | -0.04 | 0.01 | -0.20*** | 0.08** | -0.09* | 0.001 | 0.08** | -0.09** |  |
| Late Subsequent Pregnancy | -0.05 | -0.17* | -0.10 | -0.05 | -0.07 | 0.01 | -0.09** | 0.03 | -0.07 | -0.16 | 0.20*** | -0.06* | -0.08** |

Relationships among TL and epigenetic aging measures with markers of external validity were directionally consistent with prior literature; shorter TL and higher epigenetic age or faster pace of aging were generally observed in individuals with higher pre-pregnancy BMI, lower income-to-needs ratios, and greater illness burden. Correlations among these markers and TL were directionally appropriate but not significant after adjusting for maternal age. Among the epigenetic aging measures, GrimAge2 was significantly higher among individuals with higher BMI (r = 0.29, *P<*.001), greater illness burden (r = 0.27, *P<*.001), and lower income-to-needs ratios (r = –0.27, *P<*.001). PhenoAge showed similar patterns, with higher values observed among those with higher BMI (r = 0.08, *P=*.20), greater illness burden (r = 0.09, *P=*.17), and lower income-to-needs (r = –0.10, *P=*.17). DunedinPACE showed the strongest and most consistent pattern: it was positively correlated with pre-pregnancy BMI (r = 0.35, *P*<.0001) and illness burden (r = 0.28, *P*<.0001), and negatively correlated with income-to-needs (r = –0.21, *P*=.001), reinforcing its sensitivity to known social and metabolic predictors of biological aging.

### 3.2 Impact of Pregnancy and Time on Biological Aging

#### 3.2.1 TL (qPCR Telomere Length)

In the non-interaction model, significant decreases in TL were observed during Spline 2 of the time variable (*b* = –0.81, SE = 0.37, *P*=.029; **Table 3**), indicating telomere shortening later in the postpartum period. Late subsequent pregnancy was associated with significantly shorter TL (*b* = –0.79, SE = 0.36, *P*=.032), suggesting that those who became pregnant again in later follow-up had shorter TL than those that did not. No significant change was observed in the early postpartum period (Time Spline 1, *P*=.36), and an earlier subsequent pregnancy (occurring greater than 6 months outside of sampling window for biological aging measures) was not significantly associated with TL (*P*=.18). When allowing the effect of each time spline to differ by late pregnancy status the main effects weakened and a large but non-significant interaction emerged between late pregnancy status and Spline 2 (*b* = –1.27, SE = 0.97, *P*=.19), suggesting that the observed group difference in TL may reflect differential trajectories over time, rather than a static between-group difference (**Figure 1A**).

**Figure 1.**
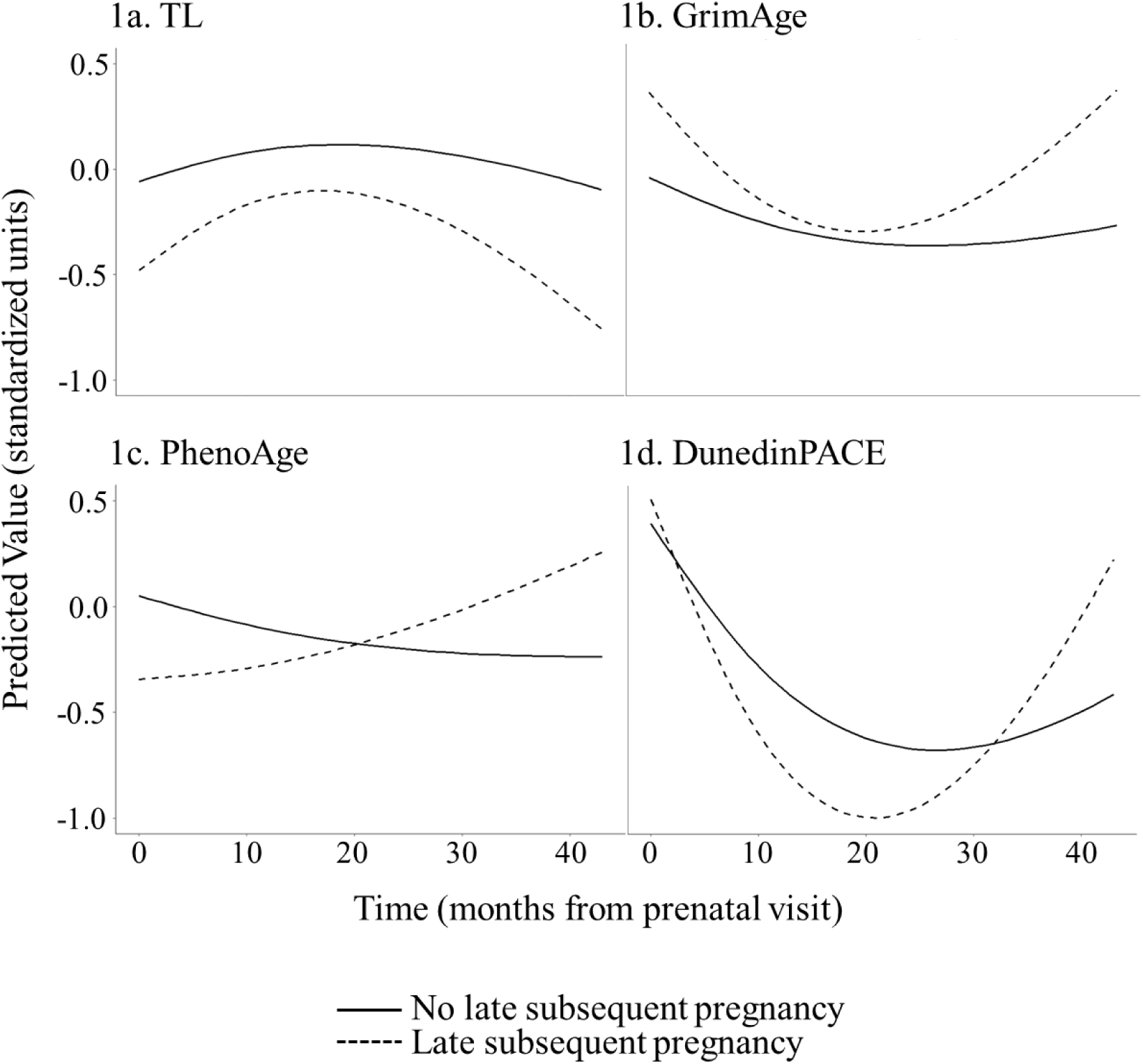
Trajectories of biological aging (predicted values in standardized units) across time by pregnancy status. All models adjusted for income-to-needs, race/ethnicity, pre-pregnancy BMI, gestational age at Time 0 prenatal visit, and illness burden; DunedinPACE also adjusted for chronological age. Trajectories are shown for (1a) absolute telomere length (TL), (1b) GrimAge2, (1c) PhenoAge, and (1d) DunedinPACE. Across most measures, pregnancy was associated with higher biological aging (shorter TL, higher epigenetic age, faster pace of aging), followed by partial recovery in the postpartum period. However, women who became pregnant again showed acceleration in biological aging, particularly evident in their pace of aging, during the time frame of the additional pregnancy.

**Table 3.** Estimates and standard errors from linear mixed-effects models predicting biological aging outcomes across time and by pregnancy status (see **Supplemental File S6** for full model results). Columns show fixed effect estimates (Estimate ± SE) from models without interaction terms (left) and models including interactions between time and pregnancy status (right). Time was modeled using natural splines to capture nonlinear changes across the postpartum period. “Late Subsequent Pregnancy” indicates whether a participant became pregnant again during the follow-up period. Significant effects are indicated with symbols: \**P<*.05, \*\**P<*.01, \*\*\**P<*.001, †*P<*.10. Outcomes include: TL (absolute telomere length), GrimAge2 and PhenoAge (epigenetic age estimators), and DunedinPACE (pace of aging estimator).

| No Interaction Models (Estimate + SE) |  |  |  | Interaction Models (Estimate + SE) |  |  |  |  |
| --- | --- | --- | --- | --- | --- | --- | --- | --- |
| Outcome | Time Spline 1 | Time Spline 2 | Late Subsequent Pregnancy | Time Spline 1<br>* Late Subsequent Pregnancy | Time Spline 2<br>* Late Subsequent Pregnancy | Time Spline 1 | Time Spline 2 | Late Subsequent Pregnancy |
| TL | 0.49 (.54) | -0.81 (.37)* | -0.55 (.42) | 0.27 (1.68) | -1.27 (.97) | 0.45 (.57) | -0.58 (.40) | -0.55 (.42) |
| GrimAge2 | -2.11 (1.06)* | 0.77 (.80) | -0.81 (.76) | -1.06 (3.80) | 2.97 (2.02) | -1.94 (1.11) | 0.21 (.87) | -0.83 (.77) |
| PhenoAge | -2.22 (2.20) | -0.16 (1.64) | -1.17 (1.51) | 5.29 (7.80) | 6.21 (4.21) | -2.47 (2.29) | -1.36 (1.81) | -1.22 (1.51) |
| Dunedin-PACE | -0.24 (.04)*** | 0.05 (.02) | -0.03 (.03) | -0.04 (.13) | 0.20 (.07)** | -0.23 (.04)*** | 0.01 (.03) | -0.03 (.03) |

#### 3.2.2 GrimAge2

In the non-interaction model, GrimAge2 acceleration declined over the early postpartum period (Time Spline 1: b = –2.11, SE = 1.06, *P*=.049), leveling off and showing non-significant change during later postpartum (Time Spline 2: b = 0.77, SE = 0.80, *P*=.33). Late subsequent pregnancy was associated with a non-significant increase in GrimAge2 (b = 1.02, SE = 0.69, *P*=.14). When allowing for time-by-group interactions, a large, though non-significant, interaction emerged in the second spline (b = 2.97, SE = 2.02, *P*=.14), consistent with faster GrimAge2 acceleration over time among those with a late subsequent pregnancy (**Figure 1B**).

#### 3.2.3 PhenoAge

In the non-interaction model predicting PhenoAge acceleration, a non-significant decrease in PhenoAge was observed over the first time spline (b = –2.22, SE = 2.20, *P*=.31), with little change in the second time spline (b = –0.16, SE = 1.64, *P*=.92), suggesting a possible early postpartum recovery pattern. Late subsequent pregnancy was not significantly associated with PhenoAge. Allowing an interaction between the Time Spline 2 and late subsequent pregnancy showed a sizable but non-significant effect in the direction of increased PhenoAge (b = 6.21, SE = 4.16, *P*=.14), suggesting a potential late-postpartum acceleration in PhenoAge for women with a subsequent pregnancy (**Figure 1C**).

#### 3.2.4 DunedinPACE

In the non-interaction model, DunedinPACE exhibited a non-linear trajectory with a significant decrease during the early postpartum period (Time Spline 1: b = –0.24, SE = 0.04, *P*<.001), indicating a slowing of the pace of aging in the initial months following pregnancy. The estimate during the later postpartum period was not significant but suggested stabilization of change (Time Spline 2: b = 0.05, SE = 0.03, *P=*.12). There was no difference in pace of aging between those with and without a late subsequent pregnancy. In the interaction model, a significant difference in trajectory emerged. The early postpartum period continued to show a significant slowing in the pace of aging (Spline 1: b = –0.23, SE = 0.04, *P*<.001), while individuals with a late subsequent pregnancy exhibited a significant increase in the pace of aging across the later time interval (Spline 2 × Late Pregnancy: b = 0.20, SE = 0.07, *P*=.006). In contrast, the reference group demonstrated relative stability across the same period. Notably, the significant interaction between Spline 2 and late subsequent pregnancy indicates that among women who became pregnant again later in the study period, the pace of aging accelerated during the time interval corresponding to the subsequent pregnancy (**Figure 1D**).

## 4. Discussion

This study investigated how biological aging shifts from pregnancy to the postpartum period and how these trajectories may be altered by an additional subsequent pregnancy. Using multiple validated biomarkers (e.g., telomere length, GrimAge2 and PhenoAge epigenetic clocks, epigenetic pace-of-aging) we modeled nonlinear within-person change across two distinct phases: an initial postpartum window shared by all participants and a subsequent period during which some women became pregnant again.

Consistent with emerging evidence, we observed that the early postpartum period (∼9 months) may serve as a window of biological recovery. For most aging measures, this phase was characterized by either no significant change or directionally favorable reductions in biological age, particularly among women who did not conceive again during follow-up. These findings closely parallel prior work showing that biological age, indexed by GrimAge2 and PhenoAge, declined from pregnancy to one year postpartum, accompanied by a rejuvenation of immune cell profiles (Ross et al., 2020). Our findings are also in line with evidence that physiological systems recover along varying timelines, with some systems, such as immune regulation, rebounding more quickly than others (Bar et al., 2025).

Our results extend these insights in important ways. We provide novel evidence that subsequent pregnancy during the recovery window may interrupt these trajectories. Across telomere length, GrimAge2, and DunedinPACE, we observed interaction effects suggesting increased biological aging in women sampled again during a subsequent pregnancy, contrasting with the stabilization or deceleration observed among women who remained postpartum. These findings align with hypotheses that additional pregnancies may carry physiological costs that could compound over time, accelerating aging beyond early postpartum recovery. For the measure of pace of aging (DunedinPACE), the interaction between time and pregnancy status was statistically robust and large enough to be potentially clinically meaningful (∼2.35 additional months of biological aging per calendar year of chronological aging), suggesting that DunedinPACE may be particularly sensitive to cumulative physiological demand and a potential indicator of longitudinal reproductive aging dynamics. The patterns emerging from our findings echo prior longitudinal work demonstrating that women who conceived again between study waves exhibited greater epigenetic age gains by approximately 2.4 - 2.8 months compared to those who did not (Ryan et al., 2024). Our findings add to this literature by providing within-person aging estimates across multiple timepoints and by directly sampling biological aging during a successive pregnancy, an approach not yet reported in previous studies.

Although measures of biological aging in our study showed directionally consistent patterns, DunedinPACE was the only measure for which the interaction term reached statistical significance. This is consistent with prior literature and may reflect greater sensitivity of DunedinPACE to systemic physiological stressors. For instance, DunedinPACE was associated with socioeconomic adversity in childhood (Raffington et al., 2021), dementia risk in adulthood (Sugden et al., 2022), and was responsive to caloric restriction (Waziry et al., 2023), while GrimAge and PhenoAge were not. These findings suggest that pace-of-aging measures like DunedinPACE may be uniquely suited to capture the biological impact of sustained or recurring physiological challenges.

Taken together, our work provides evidence of a dynamic and time-sensitive model of maternal biological aging, characterized by transient acceleration during pregnancy, partial resolution postpartum, and renewed aging strain during subsequent pregnancy. This interpretation also aligns with theoretical frameworks emphasizing the physiological costs of reproduction (Austad & Hoffman, 2018; Kirkwood, 1977). Reproduction initiates wide-ranging demands on the cardiovascular, endocrine, metabolic, and immune systems. Mechanistically, these demands may contribute to aging acceleration through several pathways: oxidative stress generated by heightened metabolic activity; inflammation from immunological adaptations to fetal tolerance; and endocrine shifts, including steep fluctuations in estrogen, progesterone, and cortisol (Shalev & Belsky, 2016). Following delivery, the body undergoes extensive remodeling, including immune resetting and shifts in hormonal and metabolic homeostasis, that may support recovery and, in some domains, confer transient rejuvenation (Bar et al., 2025). For instance, reductions in senescent T-cell populations and increases in naïve immune cells postpartum (Ross et al., 2020) have been interpreted as a temporary reversal of immunological aging. However, such recovery likely draws upon proliferative reserves, particularly in the hematopoietic system. One possibility is that the biological “reset” observed postpartum reflects the clearance of older immune cells and the appearance of younger, recently generated cells; over time, the cumulative proliferative demands of repeated pregnancies may deplete stem cell reserves, leaving women biologically older despite transient signs of postpartum recovery. This hypothesis is supported by the stepwise pattern observed in longitudinal studies: an initial pregnancy-related increase in biological age, partial postpartum reduction, and net aging gain over multiple reproductive cycles.

Our study also contributes to a broader conversation about how context may modulate biological aging responses to reproduction. Although our findings converge with previous studies in their directionality, effect sizes in our sample were modest, potentially reflecting the relatively high socioeconomic status and good health of our participants. Conducted in a high-resource setting with universal healthcare and strong social supports, a cross-sectional study of young Finnish women found no relationship between nationally registered births and four measures of epigenetic aging (Harville et al., 2021). By contrast, larger effects of pregnancy on biological aging both cross-sectionally and longitudinally have been reported in lower-resource settings, such as in the Cebu cohort in the Philippines (Ryan et al., 2024), where early and frequent childbearing occurs under greater nutritional and psychosocial stress. Evolutionary and life history models suggest that the physiological cost of reproduction may be shaped by environmental resources, maternal age, and cumulative reproductive load (Jasienska, 2009; Ziomkiewicz et al., 2016). Accordingly, the biological toll of pregnancy may be amplified in contexts of limited recovery capacity and attenuated where adequate support for physiological restoration is available.

This study offers several strengths that advance our understanding of biological aging in the perinatal period. Our longitudinal design spanned nearly four years following an initial pregnancy, one of the longest follow-up periods in this literature to date. The use of repeated within-person measurements enhanced sensitivity to change, and our multimodal assessment allowed us to triangulate across biomarkers that may reflect different aspects of the aging process. Our measures converged on similar patterns, increasing confidence in the results.

Further, our data was drawn from a cohort of women with healthy, uncomplicated pregnancies, underscoring that even in the absence of complications pregnancy represents a biologically significant challenge.

Several limitations warrant consideration. First, our biological aging measures were derived from saliva, while most aging clocks have been developed in blood. While prior work suggests moderate cross-tissue correlations (Apsley et al., 2025; Tay et al., 2025; Wolf et al., 2024), potential tissue-specific variation in biological aging signal cannot be ruled out. Second, our sample size, while sufficient for modeling within-person change, was modest by the standards of epigenetic aging studies and may have limited our ability to detect small effects. Third, although our cohort was demographically diverse, it was also relatively well-resourced. As discussed, biological aging responses to pregnancy may be more pronounced in populations exposed to greater cumulative stress or lower recovery capacity, highlighting the importance of replicating this work in other contexts. Fourth, a key limitation of our study is the absence of concurrent measures of postpartum BMI. Prior studies have shown that postpartum metabolic recovery, particularly weight loss during postpartum, is associated with greater deceleration in biological aging. For example, one study found that parity-related increases in PhenoAge were attenuated after adjusting for current BMI, suggesting that metabolic status may mediate aging trajectories (Kresovich et al., 2019). Another study reported that women who retained more weight during the one year postpartum follow-up period showed reduced reversal of epigenetic age acceleration (Ross et al., 2020). Without postpartum BMI data, we are unable to determine whether the aging patterns observed in our sample reflect the influence of pregnancy per se, or whether they were influenced by variability in metabolic status between and within individuals. It is possible that some of the between-person differences in aging trajectory were driven by differences in postpartum weight retention. However, it is notable that women who became pregnant again during late follow-up had a lower pre-pregnancy BMI coming into their initial pregnancy (Time 1) compared to those that remained postpartum (25.5 vs. 27.9; *P*=.08) which is associated with lower postpartum weight retention and a more favorable postpartum metabolic profile (Endres et al., 2015; Gore et al., 2003; Joseph et al., 2008). This suggests that our findings may reflect a conservative estimate of the average toll of pregnancy on biological aging and highlights the need for future studies that include detailed, time-sensitive postpartum metabolic data in order to disentangle these effects with greater precision.

Another important limitation of this work is the relatively low temporal resolution of biological aging measurements across the postpartum period. Although the current study spans a longer postpartum window than prior studies, the sparse measurement protocol dictated by the structure of the parent study limited our ability to capture fine-grained changes in biological aging trajectories over time. Future research aiming to characterize biological aging dynamics across pregnancy and postpartum would benefit from designs incorporating intensive repeated measures across the same developmental window. Such approaches would allow more precise modeling of short-term fluctuations, longer-term recovery patterns, and biological responses to repeated pregnancies across multiple postpartum intervals.

Relatedly, although we statistically accounted for subsequent pregnancies, our design was not able to account for the gestational age at which measurement was taken within these pregnancies. As a result, we cannot isolate the precise timing of pregnancy-induced inflection points in biological aging, nor test whether shorter interpregnancy intervals impose greater aging strain than longer ones. Future studies should incorporate serial aging measures during and between successive pregnancies, allowing for clearer resolution of when biological age acceleration emerges, peaks, and resolves.

Together, our findings provide novel longitudinal evidence that pregnancy is associated with accelerated biological aging, followed by some recovery during the postpartum period. Across multiple biomarkers and over nearly four years of follow-up, we observed consistent patterns suggesting that pregnancy imposes a substantial biological burden, even among healthy women with healthy pregnancies. These shifts in aging markers underscore the importance of understanding postpartum recovery as a dynamic, ongoing process and raise critical questions about the long-term implications of interpregnancy intervals for maternal aging. Future research should adopt a lifespan perspective, encompassing measurement from preconception through the extended postpartum period, to better understand how reproductive experiences shape biological aging trajectories and contribute to health and disease risk later in life.

## Data Availability

All data produced in the present study are available upon reasonable request to the authors.

## Notes

### Competing Interest Statement

The authors have declared no competing interest.

### Funding Statement

Work on this article was supported by a grant from the National Institutes of Health, National Institute of Nursing Research (R01 NR019610). The content is solely the responsibility of the authors and does not necessarily represent the official views of the National Institutes of Health.

### Author Declarations

The study received approval from the Institutional Review Board of Uiversity of North Carolina - Chapel Hill (#17-1914).

