## Supplemental File S1 for "Maternal Biological Aging in Mid to Late Pregnancy and across Four Years Postpartum: Evidence for Postpartum Recovery and Disruption by Subsequent Pregnancy"

Samples passing QC for study inclusion

| Time | Consented and provided saliva | Passed QC for Inclusion (%) |
| --- | --- | --- |
| 1 | 113 | 88 (78%) |
| 2 | 117 | 96 (83%) |
| 3 | 88 | 87 (99%) |

Sample size by timepoint and late pregnancy status

|  | Any | Time 0 and 6-months | 6-months and 36-months | Time 0 and 36-months | All Timepoints |
| --- | --- | --- | --- | --- | --- |
| Full Sample | 130 | 67 | 62 | 58 | 45 |
| No Late Secondary Pregnancy | 111 | 60 | 51 | 50 | 39 |
| Late Secondary Pregnancy | 19 | 7 | 11 | 8 | 6 |
