## Supplemental File S2 for "Maternal Biological Aging in Mid to Late Pregnancy and across Four Years Postpartum: Evidence for Postpartum Recovery and Disruption by Subsequent Pregnancy"

**Supplemental File S2: TRN Reporting Guidelines**

| **ITEM** | **DESCRIPTION** | |
| --- | --- | --- |
| **Sample Type, Storage, Extraction, and Integrity** | | |
| Sample type | Saliva collected via passive drool at Time 1, Time 2, and Time 3. Samples collected at Time 1 and Time 2 (n=184) were assayed together in the same batch for aTL. Samples collected at Time 3 were assayed in a second batch (n=88). | |
| DNA extraction method | DNA was extracted using QIAamp kits (Qiagen). | |
| DNA storage conditions, including freeze-thaw cycles | DNA was stored at -20°C after extraction at the Shalev Lab. On average there were four freeze-thaws for DNA samples between extraction and the qPCR assay. The first thaw was done to determine dsDNA concentration using the Quant-iT PicoGreen dsDNA assay. The second thaw was done to aliquot DNA for DNA methylation assay. The third thaw was needed to perform a dilution for the qPCR assay. The final thaw occurred when the sample was assayed for telomere length. Samples needing to be reassessed on qPCR assays (n=48; 18%) were thawed one additional time. DNA samples were stored for an average of 2.57 months between the PicoGreen assay and qPCR assay. | |
| Method of documenting DNA quality and integrity | Double stranded DNA (dsDNA) concentrations were quantified for all samples using Quant-iT PicoGreen with mean dsDNA=17.84 ng/uL. DNA purity was assessed using 260/280 and 260/230 (mean_260/280_=1.80; mean_260/230_=1.44). A subset of samples (n=27, 10%) were evaluated using the DNA Integrity Number generated by the Agilent 2200 TapeStation with mean_DIN_=2.55. No exclusionary criteria were imposed prior to assays. | |
| Percentage of samples specifically tested for DNA quality and integrity | All samples were subjected to quality control via evaluation of 260/280 and 260/230 ratios. A subset of samples were subjected to quality assessment via TapeStation. | |
| **qPCR Assay** | | |
| Method (qPCR, MMqPCR, aTL, etc.) | qPCR assays to calculate absolute telomere length (aTL) were structured such that each assay comprised two qPCR runs, one run quantifying telomere content in kilobases (T) and a second run quantifying genome copy number (S) using the single copy gene *IFNB1.* The two runs (T & S) were always performed on the same day using the same DNA aliquot which was stored at 4°C between runs (~2.5 hours). Each run hosted triplicate reactions of 22 samples, 6 standards, 5 positive controls, and 1 no template control on 100 well disks.  Time 1 and Time 2 samples were run in the first batch across a period of 11 days from 5/27/2024 to 6/06/2024. Time 3 samples were run in the second batch across a period of 12 days from 10/17/2024 to 10/28/2024 | |
| PCR machine type | Qiagen Rotor-Gene Q using 100 well disks | |
| Source of master mix and reagents, and final reaction volume | The final reaction mix for the telomeric and *IFNB1* reactions contains 1x QuantiTect SYBR Green Master Mix (Qiagen), 0.2 U Uracil Glycosylase (Thermo Fisher Scientific), 0.1 uM forward-reverse primer pair, and 6 ng DNA in a 20 uL reaction.  Primers are purchased from IDT in as pre-mixed pairs (RxnReady Primer Pool) with HPLC purification and 10uM concentration in IDTE Buffer pH 8.0. | |
| Telomere primer sequences and concentration | Forward Primer: 5'-CGG TTT GTT TGG GTT TGG GTT TGG GTT TGG GTT TGG GTT-3′  Reverse Primer: 5'-GGC TTG CCT TAC CCT TAC CCT TAC CCT TAC CCT TAC CCT-3′ | |
| Single copy gene name, primer sequences, and concentration | *IFNB1* Forward Primer: 5’-TGG CAC AAC AGG TAG TAG GCG ACA C-3’  *IFNB1* Reverse Primer: 5’-GCA CAA CAG GAG AGC AAT TTG GAG GA-3’ | |
| Full PCR program description including temperature, times, and cycle numbers | 50°C – 2min  95°C – 15min |  |
|  | 95°C – 15s | 40 cycles |
|  | 55°C for 1 min with data acquisition |  |
|  | Melt 60°C to 99°C rising 1°C per step with 5 sec per step | |
| PCR efficiency of single copy gene and telomere primers | Telo: R^2^ = 0.99828; Efficiency=1.98  IFNB1: R^2^ = 0.99657; Efficiency=1.95 | |
| Source and concentration of control samples and standard curve | 4 positive controls were selected from within the sample to control for variation across T and S runs. The final control sample was comprised of DNA extracted from the Jurkat cell line (ThermoFisher), which is known to have short telomere length (<6kb). Standards consisted of double stranded oligomers purchased from IDT as lyophilized pellet with PAGE purification.  Standard curves for T runs consisted of 84 bp double stranded oligomer comprised of 16 copies of canonical telomere repeat. Telomere Standard A had concentration 0.10 ng/uL, which equates to 5.86e+08 kb telomeric DNA when 6uL is used in the qPCR assay. A series of 1/10 serial dilutions were performed to generate a total of 6 standards for each T run comprising a range of 5.86e+08 to 5.86e+03 kb telomeric DNA.  Standard curves for S runs consisted of 83 bp double stranded oligomer corresponding to the region of IFNB1 genomic DNA flanked by IFNB1 primers. IFNB1 Standard 1 had concentration 0.00033 ng/uL, which equates to 1.18e+07 diploid genomes when 6uL is used in the qPCR assay. A series of 1/10 serial dilutions were performed to generate a total of 6 standards for each S run comprising a range of 1.18e+07 to 1.18e+02 diploid genome copies. | |
| Telomere Standard Oligomer Sequences | Sense: 5’-CCC TAA CCC TAA CCC TAA CCC TAA CCC TAA CCC TAA CCC TAA CCC TAA CCC TAA CCC TAA CCC TAA CCC TAA CCC TAA CCC TAA-3’  Anti-sense: 5’-TTA GGG TTA GGG TTA GGG TTA GGG TTA GGG TTA GGG TTA GGG TTA GGG TTA GGG TTA GGG TTA GGG TTA GGG TTA GGG TTA GGG-3’ | |
| *IFNB1* Standard Oligomer Sequences | Sense: 5-GCA CAA CAG GAG AGC AAT TTG GAG GAG ACA CTT GTT GGT CAT GTT GAC AAC ACG AAC AGT GTC GCC TAC TAC CTG TTG TGC CA-3’  Anti-sense: 5’-TGG CAC AAC AGG TAG TAG GCG ACA CTG TTC GTG TTG TCA ACA TGA CCA ACA AGT GTC TCC TCC AAA TTG CTC TCC TGT TGT GC-3’ | |
| **Data Analysis** | | |
| Mean and standard deviation or median range of telomere lengths | Across all samples:  Mean = 5.90 kb; SD = 2.04 kb.  Median = 5.90 kb; Range = 0.06 kb to 18.68 kb. | |
| Number of sample replicates | Each sample was assessed for T and S on a single run with three replicates within the run. If the sample did not pass quality control criteria described below it was run a second time. | |
| Level of independence of replicates | Replicates were drawn from the same DNA aliquot (i.e., the same tube). | |
| Analytic method, considering replicate measurements, to determine final length | Estimates of kb telomeric DNA and genome copy number were calculated automatically based on the alignment of each sample with the standard curve. When applicable, estimates for the no template control were subtracted from estimates of the analytical samples prior to calculating aTL values. The average kb telomeric DNA estimates and genome copy number estimates across triplicate measurements were used to calculate aTL values.  $aTL=\frac{Estimated kb Telomeric DNA}{Estimated Genome Copy Number\times92}$ | |
| Method of accounting for variation between replicates | When the coefficient of variation across triplicate estimates of telomere content or genome copy number was greater than 10%, replicate estimates were evaluated based upon their deviation from mean across triplicates. If the absolute distance between the estimated kb T or genome copies for a replicate and the closest replicate kb T or genome copies is greater than twice the distance between the remaining two replicates, it was considered an outlier to be excluded and the mean was recalculated using the remaining two replicates. Excepting samples that were rerun or failed, an average of 4.6 T replicates and 2.9 S replicates were dropped per run (*in this case aTL values were calculated using the average across duplicate measures*).  In the case where coefficient of variation across replicates was still greater than 10% after removal of a single outlier, or was greater than 10% without a clear outlier defined by the criteria above, the sample was reassessed for both telomere content and genome copy number, and subjected to the same quality control evaluation. | |
| Method of accounting for well position effects within plates | The unique rotary design of the Rotor Gene Q is optimized to minimize well position effects. As such no accounting for well position effects was performed. | |
| Method of accounting for between plate effects | To control for inter-assay variability, the telomeric content and genome copy number were assessed for five control samples on each T run and each S run. For each run, the estimated telomeric content and genome copy number were divided by the average estimated telomeric content and genome copy number for all runs to get a normalizing factor for that sample on a given run. This was done for all controls to get an average normalizing factor for that run. Estimates for analytical samples were then divided by the normalization factor for a given run.  This procedure was done separately for each batch. The two batches used the same set of control samples to control for inter-assay variability. The batch effect was next adjusted: first, the aTL values of the five control samples measured in the second batch were divided by the those in the first batch to derive an average normalization factor; second, all the aTL values of analytical samples from the second batch were divided by that normalization factor to get final adjusted aTL values for downstream analyses.  In this manner the average intra-run CV across replicate kb telomeric DNA estimates and genome copy number estimates was 5.32% and 4.33% respectively. The average inter-assay CV for aTL estimates of 5 control samples was 8.63% for the first batch of assay, and 9.81% for the second batch of assay. | |
| % of samples repeated and % of samples failing QC and excluding from further analyses | For the first batch,  28/184 = 15% of samples repeated  0/184 = 0 % of samples failed QC and excluded from analyses.  For the second batch,  20/88 = 23% of sample repeated  1/88 = 1% of samples failed QC and excluded from analyses. | |
| Acceptable range of PCR efficiency for single copy gene and telomere primers | 1.90 – 2.10 (5*% variation*) | |
| ICCs of samples/study groups to address variability | A selection of 20 saliva samples with repeatedly extracted DNA samples were assayed twice for the purposes of calculating the ICC. ICCs were calculated at the level of aTL values. ICC(2,1) was 0.68 and ICC(2,k) was 0.81. | |
| T/S ratio transformed to a z-score prior before comparison across methods/studies | N/A. No comparison across studies was conducted. | |
| How samples nested within families were accounted for | For the first batch, Time 1 and Time 2 samples from the same individual were run on the same plate except in cases when a single sample from a given individual needed to be rerun due to high intra-assay CV. | |
