## Supplemental File S3 for "Maternal Biological Aging in Mid to Late Pregnancy and across Four Years Postpartum: Evidence for Postpartum Recovery and Disruption by Subsequent Pregnancy"

**Supplemental File S3.** Cell Heterogeneity

The Houseman-ReferenceFree method can be used for any type of tissue (including non-blood tissue) and will provide estimates for cellular proportions based on DNA methylation data and a user-defined input that specifies the number of cell types to estimate (Houseman et al. 2016). In addition to providing cellular proportions, the Houseman-ReferenceFree method also provides a protocol for estimating the “best-fitting” number of cell subtypes in a provided sample. Briefly, deviance statistics are computed for each possible number of cell types present (2 cell types, 3 cell types, … ). These deviance statistics are computed for a random selection of the study sample a defined number of times (we used 500 bootstrapped samples). Then, summary statistics for each possible number of cell types present are compared and the number of cell types with the minimum deviance summary statistic are chosen. We performed these calculations with our buccal and saliva samples separately. We extracted the 10,000 probes with the highest variation in buccal and saliva separately and subsequently used these probes in cellular composition estimates. After computation, we determined that saliva tissue contained 7 cellular subtypes.
