## Supplementary figures and images for "Maternal Biological Aging in Mid to Late Pregnancy and across Four Years Postpartum: Evidence for Postpartum Recovery and Disruption by Subsequent Pregnancy"

### Supplemental File S4

**Supplemental File S4.** Original TL distribution

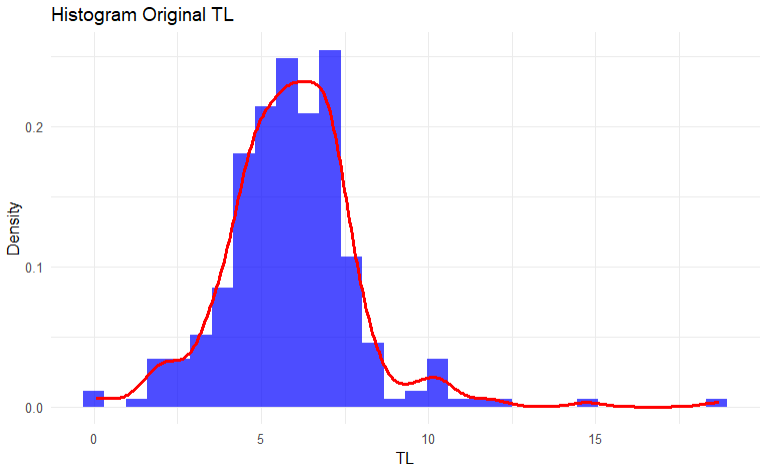

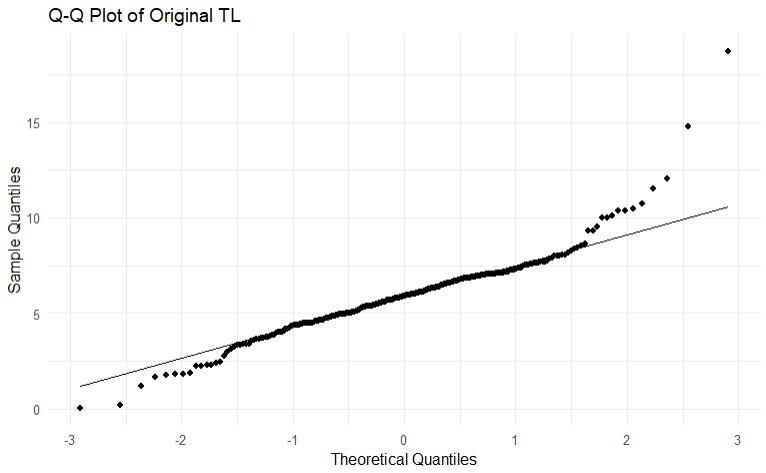
